# Distinct Relaxation Kurtosis Imaging-Derived White Matter Microstructural Phenotypes Tracking Alzheimer’s Disease Pathology and Cognitive Decline

**DOI:** 10.64898/2026.11.17.26368344

**Authors:** Dmitriy A. Yablonskiy, Satya V.V.N. Kothapalli, Cihat Eldeniz, John C. Morris, Tammie L.S. Benzinger

**Affiliations:** Department of Radiology, Washington University in St. Louis, St. Louis, MO 63110, USA; Knight Alzheimer Disease Research Center, Washington University in St. Louis, St. Louis, MO 63110, USA; The Hope Center for Neurological Disorders, Washington University in St. Louis, St. Louis, MO, USA; Department of Neurology, Washington University in St. Louis, St. Louis, MO 63110, USA

## Abstract

White matter (WM) pathology is increasingly recognized as an early, mechanistically relevant feature of Alzheimer’s disease (AD), yet sensitive in-vivo biomarkers remain limited. Here, we introduce an MRI-based Relaxation Kurtosis Imaging (RKI) technique sensitive to WM microstructure. RKI metrics differentiate two distinct phenotypes of white matter that follow divergent microstructural trajectories in preclinical and early symptomatic AD. We examined their relationships to AD progression, gray matter (GM) pathology, and clinical impairment in 305 adults spanning amyloid-negative to amyloid-positive stages with normal cognition, preclinical AD, and early symptomatic AD. Pathological changes in one phenotype reflected axonal alterations without myelin loss and were anatomically coupled to GM cortical pathology, strengthening with disease stage – a signature of Wallerian degeneration. Alterations in the other phenotype represented WM demyelination. A model incorporating RKI-metrics, age, sex, amyloid status, APOE genotype, and brain volume classified Clinical Dementia Rating (CDR)-based impairment with an area under the receiver operating curve (AUC) = 0.933, with the RKI-metrics contributing 27% (second only to amyloid at 40%). While both phenotypes related comparably to concurrent cognition (each ∼27%), the “axonal” phenotype showed the strongest relationship to subsequent cognitive decline (41% of explained variance), with the “myelin” phenotype contributing 31%. These RKI-defined dual white matter phenotypes provide new insight into AD pathology, adding diagnostic and prognostic value for tracking both current cognitive status and future cognitive decline.

**Brief Summary:** A new MRI-based Relaxation Kurtosis Imaging (RKI) method characterizes two distinct white matter tissue signatures that track Alzheimer’s progression, predicting both clinical dementia stage and future cognitive decline

**Highlights:**

- Relaxation Kurtosis Imaging (RKI) differentiates two distinct microstructural phenotypes with different trajectories of white matter pathology in Alzheimer’s disease.
- The first phenotype highlights microstructural axonal alterations anatomically coupled to regional cortical pathology consistent with Wallerian neurodegeneration.
- The second phenotype represents demyelination diffusely connected to cortical pathology.
- RKI metrics track AD pathology and predict future cognitive decline.

## INTRODUCTION

Alzheimer’s disease (AD) is defined pathologically by amyloid-beta plaques and neurofibrillary tau tangles, and the great majority of AD biomarker development has followed directly from this two-pathology framework (*1*). White matter, by contrast, has historically been treated as a secondary casualty of gray matter degeneration. This view has been substantially revised over the past two decades, as postmortem, animal-model, and neuroimaging studies have converged on white matter and myelin pathology as an early, and possibly upstream, feature of AD, capable of interacting bidirectionally with amyloid and tau rather than merely following their accumulation (*2, 3*). Recent reviews have explicitly framed this as a paradigm shift, from viewing white matter degeneration as a passive consequence of neurodegeneration to recognizing it as an active regulator of disease progression, with microstructural white matter change reported to precede gray matter atrophy and cognitive decline (*4*). Oligodendrocytes and their precursors, the cells responsible for producing and maintaining myelin, are impaired early in AD, with amyloid-beta and tau pathology disrupting oligodendrocyte precursor cell differentiation and inducing cellular senescence, further exacerbating neuroinflammation and reducing the brain’s capacity for remyelination (*5*).

Several independent lines of evidence support an early, mechanistically active role for myelin dysfunction. Myelin water fraction, an MRI measure of myelin content, is reduced in preclinical AD and associated with cerebrospinal fluid biomarkers of amyloid and tau pathology before overt cognitive symptoms (*6*). A large ADNI analysis further found this relationship is temporally structured: early white matter changes preferentially track subsequent amyloid accumulation, while later changes track tau, consistent with demyelination evolving alongside, rather than simply following, the amyloid-tau cascade (*7*). Another recent diffusion MRI study similarly found that axonal degeneration is detectable already in mild cognitive impairment and tracks the rate of subsequent clinical decline, matching established amyloid and tau biomarkers in predicting future impairment (*8*). Consistent with a preclinical role for this interaction, white matter hyperintensity volume correlates longitudinally with amyloid PET, structural MRI, and cognition, with accelerated change beginning around age 60 (*9*) and CSF proteomic profiles associated with white matter diffusion metrics differ by amyloid status in cognitively normal older adults, with amyloid-positive individuals showing distinct protein associations involving axonogenesis and synaptic organization pathways (*10*). Mechanistically, AD mouse models show that genetically induced demyelinating injury actively drives amyloid deposition, by causing amyloid precursor protein-cleaving machinery to accumulate within damaged axons and by diverting disease-associated microglia away from their normal plaque-clearing role (*11*). Single-cell sequencing studies have identified a disease-associated oligodendrocyte subpopulation in AD mouse models and postmortem human brain, in which aberrant intracellular signaling impairs axonal myelination; experimentally reversing this signaling defect ameliorates both amyloid pathology and cognitive decline in mice (*12*), a finding consistent with broader single-cell evidence implicating oligodendrocyte dysfunction across human brain disorders (*13*). Beyond amyloid-producing mechanisms, microglia are increasingly recognized as central regulators of myelin maintenance and regeneration throughout life, with myelin damage occurring both in normal aging and across neurodegenerative diseases including AD (*14*). Efforts to model these processes using human induced pluripotent stem cell-derived brain systems, including myelination itself, reflect the field’s growing recognition of white matter’s disease relevance (*15*). The prolonged, evolutionarily recent process of human myelination was proposed to render late-myelinating association white matter uniquely vulnerable to age- and disease-related breakdown, with resulting myelin damage potentially promoting amyloid deposition rather than only following it (*16*). This body of work motivates white matter status as a target for quantitative, in vivo AD biomarker development, complementary to amyloid- and tau-based measures.

Existing white matter biomarker approaches illustrate both the promise and the current limitations of this need. Diffusion analysis has revealed tau-related white matter changes in early AD that are separable from those driven by amyloid (*17*), and from those attributable to coexisting cerebral small vessel disease specifically (*18*). Diffusion-based neuroinflammation imaging correlates with amyloid status and predicts longitudinal cognitive decline, indicating that white matter inflammatory processes carry prognostic information beyond diffusion microstructure alone (*19*). Quantitative susceptibility mapping applied to brain imaging has linked magnetic susceptibility - itself sensitive to both iron and myelin content - to genetic variants involved in myelin biology (*20*). Report based on quantitative Gradient Recalled Echo (qGRE) MRI (*21*) identified significantly lower gray matter neuronal density and white matter myelination in people with amyloid level below the conventional amyloid-positive threshold despite the absence of atrophy (*22*).

In this paper we build upon the traditional qGRE approach (*21*) that relies on biophysical model of GRE signal allowing isolation of a tissue-specific transverse relaxation rate constant, R2t*, from mesoscopic magnetic susceptibility effects, vascular BOLD (blood oxygen level dependent) and macroscopic field inhomogeneity contributions to the conventional R2* relaxation signal. Using gene expression data from the Allen Human Brain Atlas, the R2t* component of qGRE signal has been shown to reflect genetically defined cellular composition of brain tissue, thus validating its biological interpretability (*23*). This framework has demonstrated sensitivity to tissue-level pathology in multiple sclerosis (*24*), traumatic brain injury (*25*) and AD (*26, 27*). The present work expands the qGRE approach by introducing R2t*-based Relaxation Kurtosis Imaging (RKI) accounting for tissue microstructural heterogeneity. In white matter RKI distinguishes two tissue phenotypes: one, where K-metric is positive due to domination of the microstructural relaxation variability (henceforth called Kr), and another, where K-metric is negative due to domination of the microstructural frequency variability (henceforth called Kf). These two phenotypes exist in the healthy control population, but they relate to clinical and cognitive status through partially distinct pathways and carry largely independent diagnostic and prognostic information.

This paper characterizes this dual phenotype’s relationship to Alzheimer’s disease progression, its anatomical coupling to local GM pathology, its value in classifying clinical cognitive impairment, and its relationship to cross-sectional and longitudinal cognitive performance, revealing consistent, generalizable disease signatures.

## RESULTS

The study cohort comprised 314 older adults recruited through the Washington University Knight Alzheimer Disease Research Center, spanning cognitively normal to mildly clinically impaired status defined by the Clinical Dementia Rating® (CDR®) (*28, 29*), and including both amyloid-positive and amyloid-negative individuals as well as both APOE ε4 carriers and non-carriers. The demographic information along with APOE, Amyloid status and CDR are detailed in Table 1. For convenience and completeness of presentation, the information is organized in 7 groups. Note that information on the tau pathology status was not available for the whole cohort, hence not included in the analysis.

**Table 1.** Demographic and clinical characteristics of study participants.

| **Group** | **Definition** | **n** | **Age, years (mean ± SD)** | **Female, %** | **APOE ε4** | **Amyloid** | **CDR (median [range])** |
|---|---|---|---|---|---|---|---|
| G1 | Unimpaired, non-carrier | 128 | 69.4 ± 7.6 | 57.0 | Non-carrier | Negative | 0 [0–0] |
| G2 | Unimpaired, carrier | 63 | 64.8 ± 8.6 | 52.4 | Carrier | Negative | 0 [0–0] |
| G3 | Preclinical, non-carrier | 32 | 74.1 ± 6.0 | 56.2 | Non-carrier | Positive | 0 [0–0] |
| G4 | Preclinical, carrier | 51 | 70.6 ± 6.6 | 60.8 | Carrier | Positive | 0 [0–0] |
| G5 | Impaired, non-carrier | 9 | 73.9 ± 9.6 | 33.3 | Non-carrier | Positive | 0.5 [0.5–1.0] |
| G6 | Impaired, carrier | 22 | 74.7 ± 6.0 | 45.5 | Carrier | Positive | 0.5 [0.5–1.0] |
| G7 | Discordant (excluded) | 9 | 72.8 ± 6.9 | 22.2 | Non-carrier | Negative | 0.5 [0.5–1.0] |

### RKI Model and Parameters’ Distribution Across WM

Quantitative gradient recalled echo (qGRE) MRI (*21*) is based on gradient-recalled-echo sequence with multiple gradient echoes (mGRE) and conventionally characterizes each brain region by a single value of R2t*, a transverse relaxation rate parameter specific to brain tissue itself, obtained after removing the contribution of the vascular BOLD effect (*30*) and magnetic field inhomogeneities (*31*). This standard approach is valid if microscopic heterogeneity in local distribution of R2t* relaxation rate parameters averages out during signal acquisition. In RKI approach, we relax this assumption by examining the microstructural distribution of R2t* and frequency values within each region, rather than only their mean values, using kurtosis, a statistical measure of how far a distribution departs from a simple, single-valued shape (see details in the Methods section).

Two physically distinct sources contribute to this kurtosis with opposite sign: variability in local tissue relaxation rates contributes positively, while variability in local frequency shifts, which causes signal dephasing among differently shifted water populations, contributes negatively. In gray matter, the frequency contribution usually dominates, yielding a net negative kurtosis. Applying this same analysis to white matter, we discovered that RKI separates WM tissue into two distinct populations according to which source of heterogeneity dominates: a population with net positive kurtosis metric, which we term Kr, in which relaxation-rate variability dominates, and a population with net negative kurtosis metric, which we term Kf, in which frequency-shift variability dominates. This partition emerged empirically from the data reflecting the multilamellar myelin sheath’s tendency to form multiple, incompletely exchanging water compartments (intra-axonal, myelin, and extracellular) within white matter, whose relative contribution to relaxation versus frequency heterogeneity can differ from location to location. Kr and Kf therefore each quantify the degree of microstructural heterogeneity - how variable the local relaxation and frequency environment is - within their respective tissue population. Applying this same positive/negative partition to the corresponding microstructural R2t* values, yields two further parameters, krR2t* and kfR2t*: the mean microstructural R2t* values (denoted here as kR2t* with “k” added to distinguish the kurtosis-model-defined mean R2t* value) computed separately within the Kr-dominated (krR2t*) and Kf-dominated (kfR2t*) tissue populations. This new set of parameters (Kr, Kf, krR2t* and kfR2t*) bear new information on WM tissue microstructure, beyond a single R2t* value. Full mathematical details of the RKI approach are provided in Materials and Methods section.

Example of RKI quantitative maps in WM is shown in Fig. 1, while kurtosis parameters’ (K and kR2t*) distribution in healthy control group (G1, see Table 1) across 41 FreeSurfer regions of interests (ROIs) is shown in Fig. 2. While RKI identified visible variability in WM kurtosis metrics (K and kR2t*), a similar contrast is not obvious on the standard MPRAGE and FLAIR images (Fig. 1).

**Figure 1.**
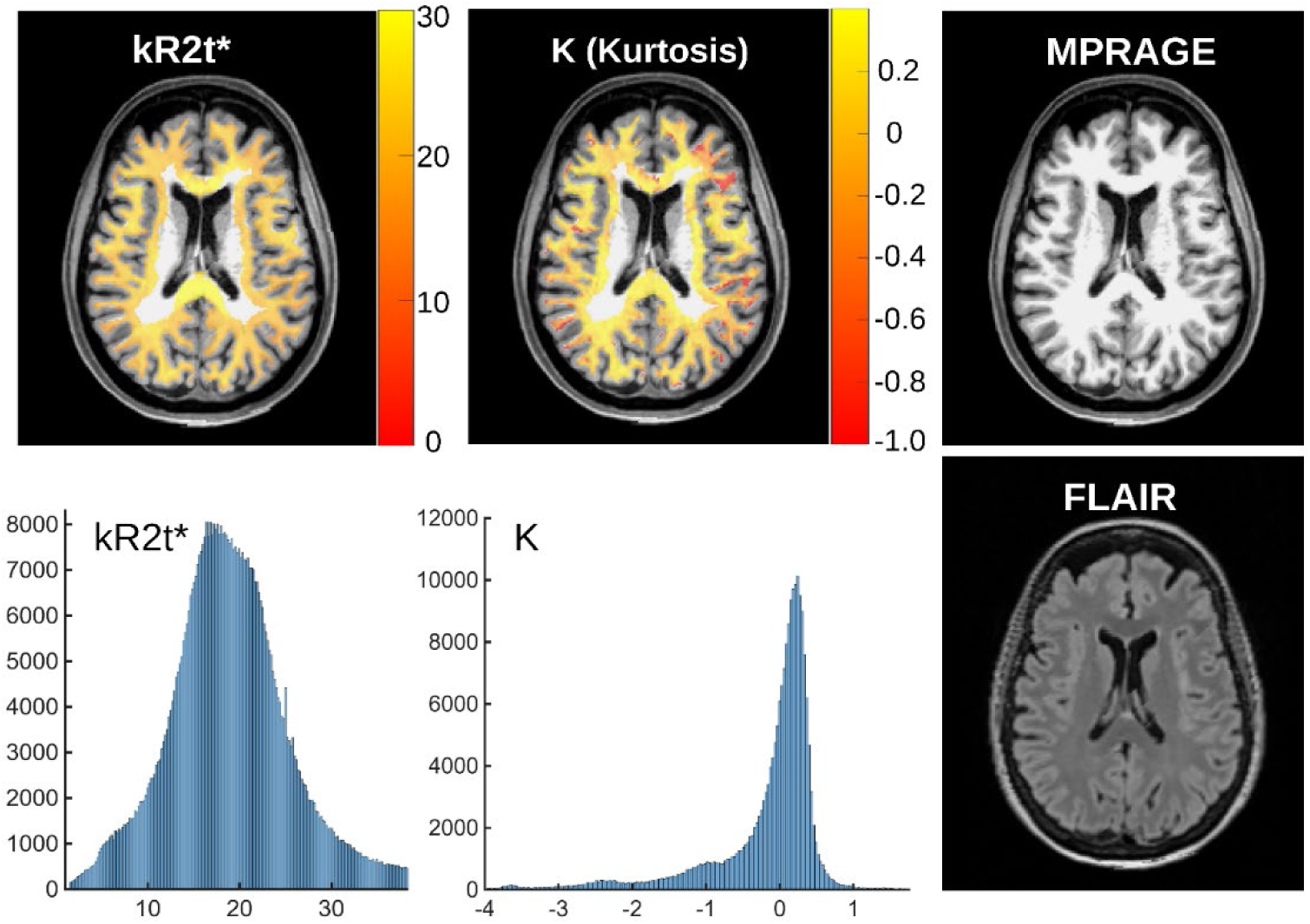
Example of representative white matter RKI maps of K (kurtosis) and kR2t* (mean microstructural R2t* value) in a single participant. K and kR2t* values, restricted to voxels within the white matter mask, are superimposed on the corresponding MPRAGE image for anatomical reference; FLAIR is shown for the same slice. Histograms show the distribution of K and kR2t* values across all white matter voxels in this participant’s brain. K values span both positive and negative ranges, reflecting the two dissociable tissue populations (Kr and Kf) characterized throughout this study.

**Figure 2.**
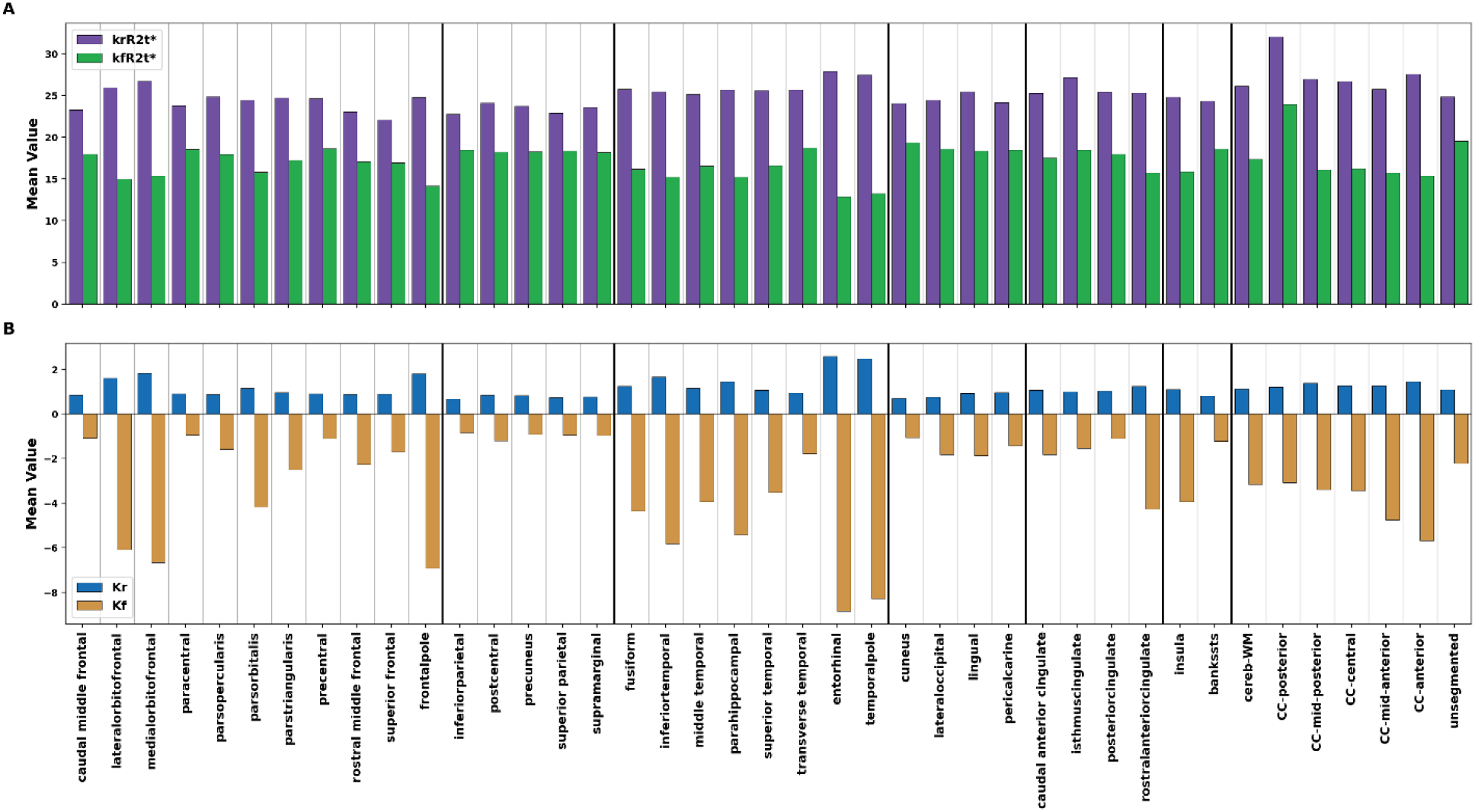
The four RKI model parameters across all 41 WM ROIs, healthy controls. Upper panel: krR2t* and kfR2t* - the mean R2t* values within the positive and negative kurtosis phenotypes. Lower panel: Kr and Kf, the underlying positive and negative kurtosis values. Regions grouped by lobe; vertical lines separate individual ROIs (light gray) and lobe groups (dark). Data represents mean values averaged across similar ROIs in right and left hemispheres.

Across the 41 established white matter ROIs examined in healthy controls (G1), Kf’s magnitude consistently exceeds Kr’s, most prominently in entorhinal and temporal pole white matter (Fig. 2, lower panel). The krR2t* and kfR2t* show the mirrored pattern where krR2t* exceeds kfR2t* across the entire white matter (Fig. 2, upper panel).

### Kr and Kf increase with amyloid-related disease progression

To assess disease-stage progression with adequate statistical power at the impaired stage (where the non-carrier pathway includes only 9 subjects), subjects from both APOE ε4 pathways were pooled at each disease stage. Each subject’s value was first age-adjusted using their own pathway’s healthy reference group (non-carrier or carrier), then expressed as a deviation from that reference group’s mean values. This yielded three pooled disease-stage groups: Unimpaired (n=191), Preclinical (n=83), and Impaired (n=31).

RKI metrics Kr and Kf both increased in magnitude with disease progression, in opposite directions (endpoint P<0.001 for both) (Fig. 3A). krR2t* also increased significantly with disease stage (endpoint P=0.002), while kfR2t* did not reach significance (endpoint P=0.090) (Fig. 3B). Notably, both Kr and Kf also reached significance in deep WM, the FreeSurfer-segmented WM lying below the 5-mm subcortical WM band, with the same opposite-direction pattern as the subcortical result (endpoint P<0.001 and P=0.002, respectively) (Fig. 3C), indicating that the disease-related changes captured by these kurtosis-derived metrics are not confined to subcortical WM but extend into deeper white matter. krR2t* and kfR2t* did not reach significance in this region (P=0.12 and P=0.28, respectively).

**Figure 3.**
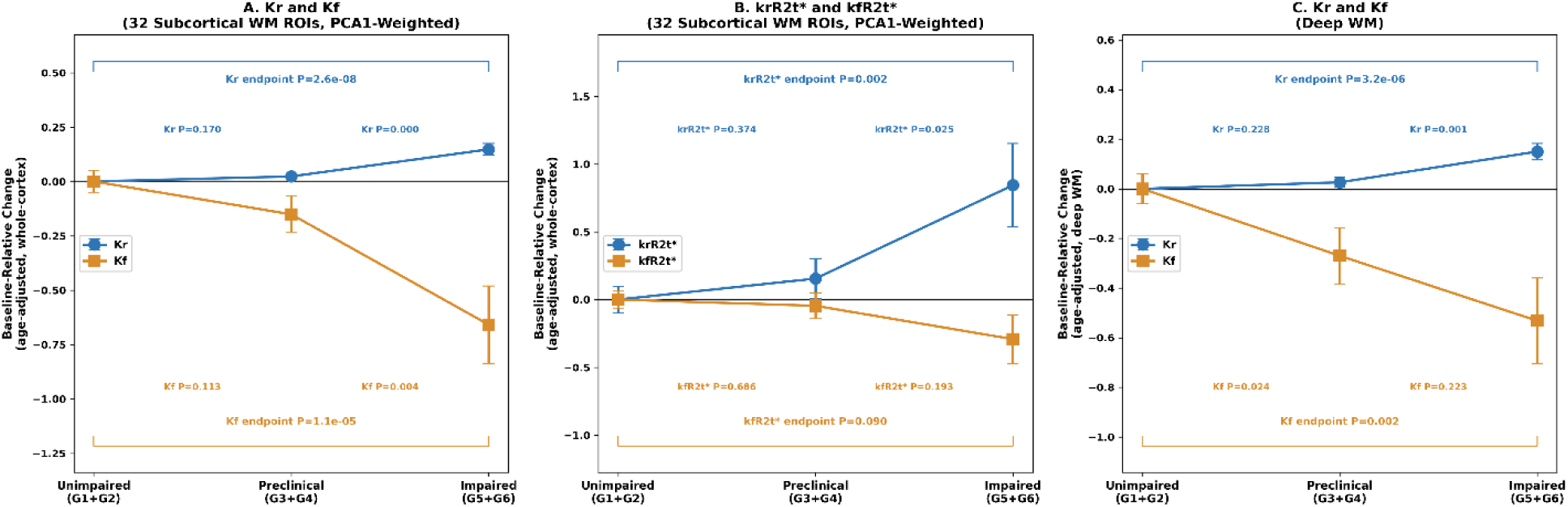
Disease-progression curves for RKI metrics Kr, Kf, krR2t*, and kfR2t*. In A (Kr, Kf) and B (krR2t*, kfR2t*) each metric is a single PCA1-weighted score summarizing the 32 established subcortical white matter ROIs per participant (see Methods), pooled across APOE ε4 pathways via age-adjusted, baseline-relative centering (see Methods). In C (Kr, Kf) are within deep WM — the FreeSurfer-segmented WM lying below the 5-mm subcortical WM band used in panels A-B, not assigned to any individual cortically-matched ROI — shown separately to demonstrate that these effects extend beyond subcortical WM. The p values for adjacent-stage comparisons are shown above/below the connecting line; p values for the endpoint (Unimpaired-vs-Impaired) comparison are shown with full-width brackets. Error bars represent standard errors of the mean values (SEM).

### Kr’s coupling to gray matter pathology strengthens with disease stage

We next tested whether each white matter parameter’s value in a given region tracks gray matter pathology in the anatomically corresponding cortical region (matched pairs defined by FreeSurfer segmentation) more closely than in other, unrelated regions (remote pairs, averaged). Gray matter pathology was quantified as dark matter (DM) Fraction (*27*), the fraction of a cortical region’s voxels with low R2t*, a measure derived from the same qGRE acquisition in GM. Herein the DM status was defined relative to a healthy reference distribution built from a subset of cognitively unimpaired participants (age <64 years, APOE ε4-negative, and cortical amyloid standardized uptake value ratio <1.42). For each FreeSurfer-defined cortical region, a pooled histogram of R2t* across these reference participants was constructed, and voxels with R2t* values below (mean - 1.96×SD) of this reference distribution were classified as DM. DM Fraction for a given participant and region is the proportion of that region’s voxels meeting this criterion. Coupling between each white matter parameter and DM Fraction was computed as the correlation between each of 32 cortical gray matter regions’ DM Fraction and the corresponding white matter ROI’s RKI parameter value, separately for matched and remote pairs, using the same baseline-relative pooling described above.

In matched pairs (Fig. 4A), Kf’s coupling to gray matter DM was strong (∼ −0.6) and stable across disease stages; kfR2t* closely tracked Kf. krR2t*’s coupling was close to zero at every disease stage. Kr’s coupling was more modest in magnitude (∼0.3) and increased with disease stage, from 0.30 (Unimpaired) to 0.34 (Preclinical) to 0.37 (Impaired). Comparing matched to remote pairs (Fig. 4B), Kr’s matched-pair coupling exceeded its remote-pair coupling at every disease stage, and this matched-versus-remote difference widened at the impaired stage; Kf, krR2t*, and kfR2t* showed comparable matched-versus-remote patterns to one another, without a comparable disease-stage-dependent widening.

**Figure 4.**
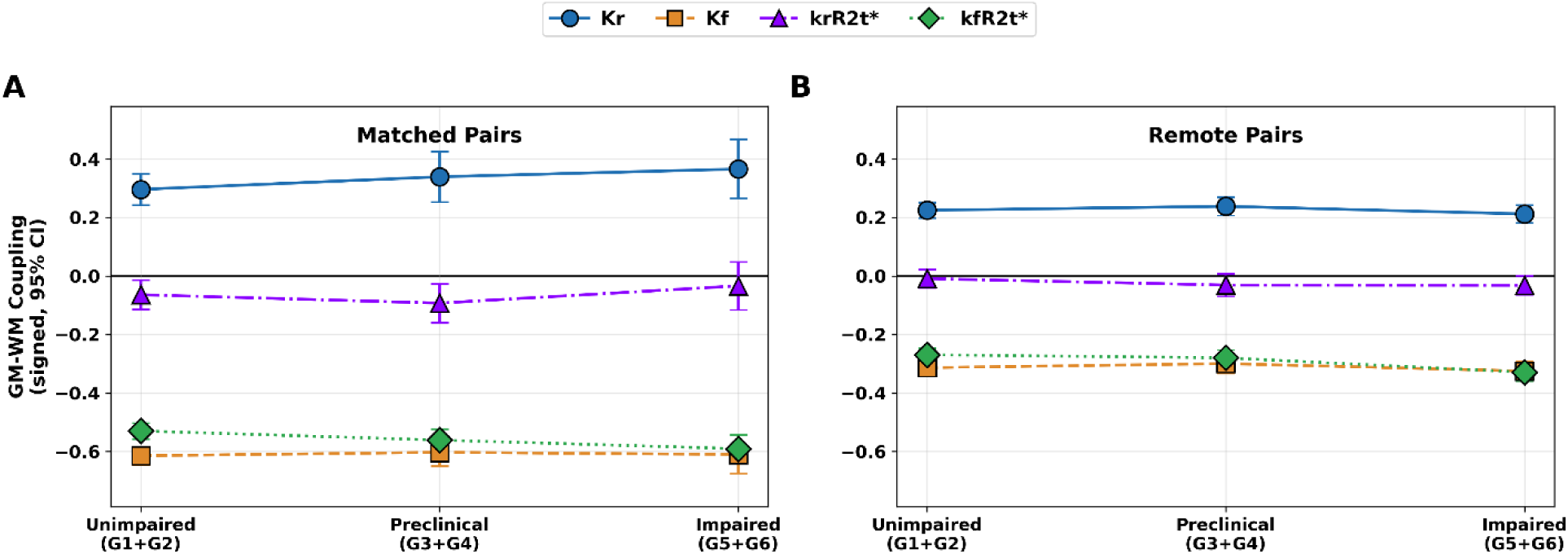
Coupling of RKI metrics Kr, Kf, krR2t*, and kfR2t* with DM Fraction (gray matter pathology). Data show signed correlations, pooled across APOE ε4 pathways. Left panel: Matched (anatomically corresponding) gray-white matter pairs. Right panel: Remote (non-corresponding) pairs, averaged. Both panels share the same vertical scale. Error bars show 95% CI across the 32 ROIs. G1-G6 groups are identified in Table 1.

### Kr metric adds diagnostic value beyond amyloid status and APOE genotype

We tested diagnostic value of Kr for classifying clinical impairment (CDR-positive vs. CDR=0) beyond established clinical and genetic predictors, using repeated 10-fold cross-validation (n=305). Kr was entered as a single whole subcortical WM score per participant, computed by weighting each of the 32 regional Kr values by that region’s loading on the first principal component of Kr variation in healthy controls (PC1 explained 50.5% of variance). A model combining Kr with age, amyloid status, and APOE genotype achieved CV-AUC=0.925; adding brain volume improved this modestly to CV-AUC=0.929. Further adding sex to this model improved performance to CV-AUC=0.933 (Fig. 5A). In Shapley decomposition of this best-performing model (Kr+Age+Amyloid+APOE+Brain Volume+Sex), amyloid status contributed most to the overall model performance (40.0%), followed by Kr (27.3%); age, APOE genotype, brain volume, and sex contributed the remainder (Fig. 5B).

**Figure 5.**
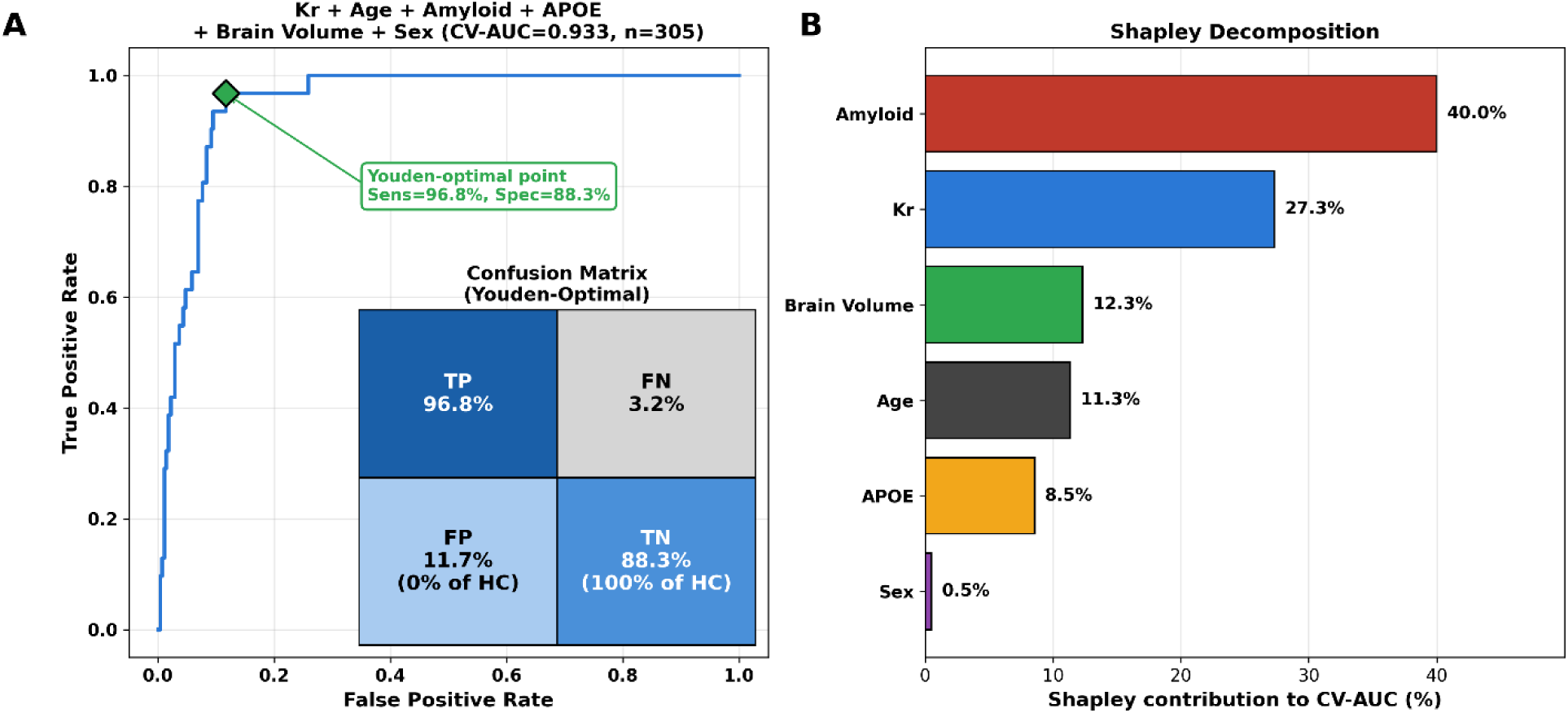
ROC curve and Shapley decomposition for the best-performing model (Kr + Age + Amyloid + APOE + Brain Volume + Sex; n=305; CV-AUC=0.933). Diamond marks the Youden-optimal operating point (sensitivity=96.8%, specificity=88.3%, decision threshold=0.191). The inset shows the confusion matrix (True Positive (TP), False Negative (FN). False Positive (FP), and True Negative (TN) results) at this threshold, with the percentage of the healthy control group (n=128) falling into each CDR=0 outcome cell shown in parentheses.

To assess whether the other three RKI model parameters, or DM Fraction (a measure of gray matter pathology; see above), provide comparable diagnostic value, each was substituted for Kr in turn within this same model (Age+Amyloid+APOE+Sex+Brain Volume+[parameter], n=305 for each). This produced a graded pattern: Kf gave a smaller but appreciable improvement over the covariates-only model (CV-AUC=0.920 vs. 0.908 without any kurtosis-model or gray matter parameter), while krR2t*, kfR2t*, and DM Fraction each gave minimal improvement (CV-AUC=0.913, 0.910, and 0.909, respectively), each within the range of cross-validation variability of the covariates-only model.

At the Youden-optimal threshold shown in Fig. 5, sensitivity was 96.8% (missing 1 of 31 CDR-positive participants), and specificity was 88.3%. All 128 healthy-control participants were correctly classified as true negatives.

### Kr and Kf metrics relate to current cognition; only Kr predicts decline

We examined how each white matter RKI parameter relates to cognitive performance, both current performance and its subsequent rate of change, using the Global Composite cognitive score. Current cognitive performance was assessed using the closest available assessment to each participant’s MRI scan within a one-year window (mean interval 94.5 days, median 84.5 days, interquartile range 52-125 days). Two joint-regression models were tested, each with age, amyloid status, APOE genotype, and sex as covariates and one of five RKI/DM parameters (Kr, Kf, krR2t*, kfR2t*, or DM Fraction) entered as a fifth simultaneous predictor: (A) current Global Composite score as the outcome (n=305), and (B) the subsequent rate of Global Composite change, a per-subject linear rate computed from cognitive assessments obtained after the MRI scan and spanning at least one year, as the outcome (n=128; mean follow-up span 3.3 years, median 2.4 years, interquartile range 1.4-4.6 years; median 3 assessments per participant, range 2-12). Each model’s total R^2^ was decomposed into per-predictor Shapley values; the adjacent narrow panel in each case shows the simple, unadjusted Pearson correlation (r) between the RKI/DM parameter and the outcome alone.

For current Global Composite score (Fig. 6A), Kf and Kr were the two strongest and most significant predictors, with comparable Shapley shares (Kf: 27% of R^2^=0.182; Kr: 27% of R^2^=0.172) and comparable simple correlations (r=+0.27 and r=-0.26, respectively). kfR2t* showed a smaller but significant contribution (12% of R^2^=0.164); krR2t* and DM Fraction did not contribute significantly to this model.

**Figure 6.**
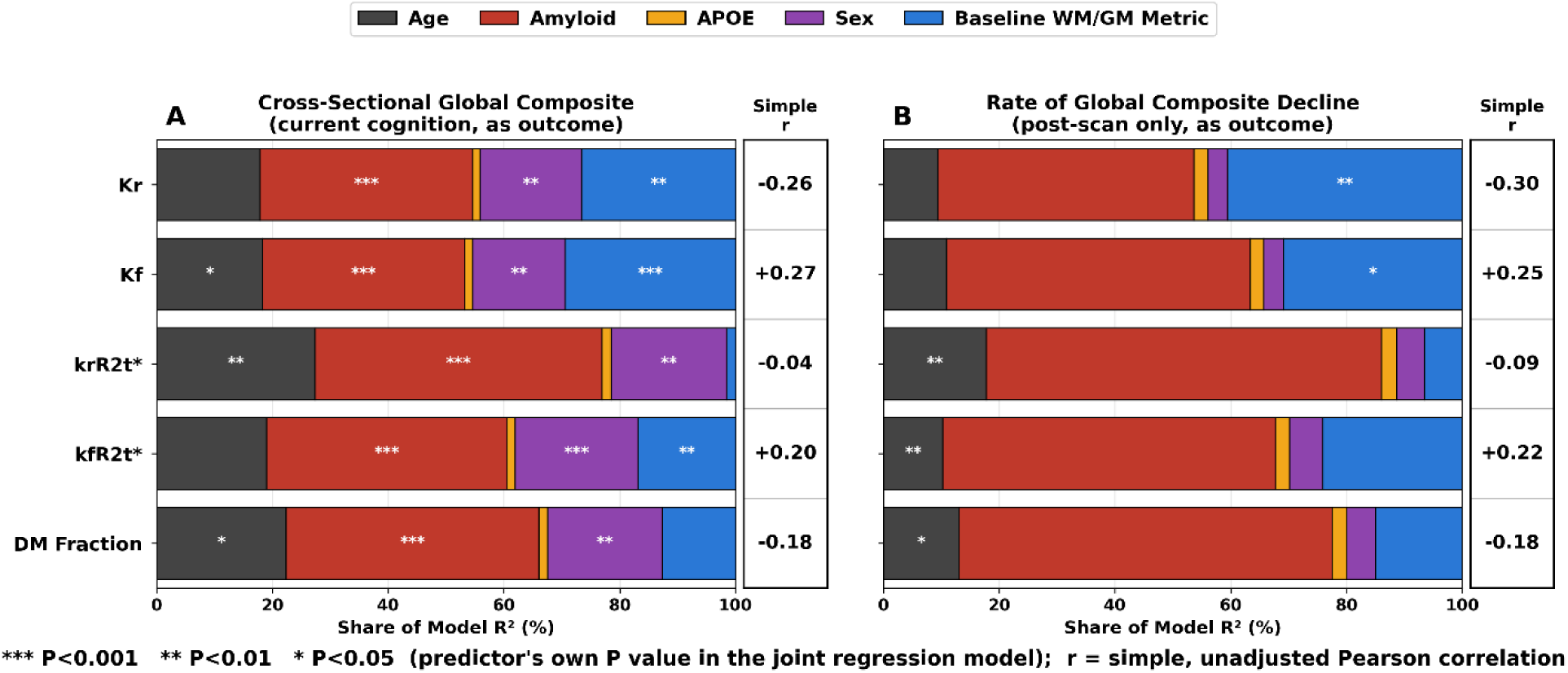
Shapley decomposition of R^2^ for Global Composite, cross-sectional (A) and as a subsequent, post-scan rate of decline (B), for each of five candidate RKI/DM parameters with age, amyloid status, APOE genotype, and sex as covariates. Significance markers denote each predictor’s own P value in the joint regression model (***P<0.001, **P<0.01, *P<0.05). The narrow right-hand panel in each case shows the simple, unadjusted Pearson r between the parameter and the outcome alone. n=305 (A), n=128 (B).

For the subsequent, post-scan rate of Global Composite decline (Fig. 6B), Kr’s Shapley contribution was the largest of any RKI/DM parameter (41% of that model’s total R^2^=0.160, P=0.0099); Kf also reached significance with a smaller contribution (31% of R^2^=0.141, P=0.045). krR2t*, kfR2t*, and DM Fraction did not contribute significantly to this model.

## DISCUSSION

This study introduces an MRI-based Relaxation Kurtosis Imaging (RKI) technique for studying microstructure of biological tissues. RKI is based on mGRE pulse sequence (available from most manufacturers) and theoretical biophysical kurtosis model (Eqs. [1]-[12] in the Methods section) connecting mGRE signal with tissue microstructural parameters. The RKI technique provides information on biological tissue microstructure beyond the commonly used mean values of GRE signal relaxation rate parameters and signal frequencies by also accounting for variability (heterogeneity) of biological water distribution among the underlying cellular matrix components. This variability is accounted for in RKI approach by means of the kurtosis metric K (Eqs. [2] and [5]) that has contributions from two distinct tissue microstructural characteristics - variances of relaxation rate parameters R2t* (Eq. [6]), and variances of inter-component frequencies (Eq. [7]). The important feature of the K metric is the opposite sign contribution of the relaxation (positive) and the frequency (negative) variances. Accordingly, we call kurtosis K in the brain regions with positive kurtosis (relaxation dominated) as Kr, while in the brain regions with negative kurtosis (frequency dominated) as Kf. The corresponding mean values of the relaxation rate parameters in these regions are identified as krR2t* and kfR2t*.

Result of RKI analysis in study human participants demonstrated the presence of WM regions with positive (Kr) and negative (Kf) kurtosis metrics. We distinguish these regions as white matter phenotypes because they play distinct roles in Alzheimer’s disease. The Kr and Kf metrics in these two WM phenotypes change in opposite directions with amyloid-related disease progression, coupled to gray matter pathology through distinct anatomical patterns, and carrying markedly different diagnostic and prognostic values. The kurtosis sign also reflects a baseline anatomical distinction between two WM phenotypes, separate from the disease-related changes. The regions with the largest Kf magnitude (entorhinal, temporal pole, and other frontal/temporal polar and orbital white matter, Fig. 2, lower panel) correspond closely to the narrow rim of superficial white matter (SWM) beneath the cortex, shown by 7T susceptibility imaging and histology to contain elevated iron without a corresponding increase in myelin content (*32*). Most Kf-dominated ROIs are gray-matter-adjacent, but not all adjacent ROIs belong to Kf phenotype which might be explained by insufficient baseline iron to overcome variance of 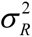 (Eq.[9]). Notably, both Kr and Kf also showed significant disease-stage changes in the deep WM region lying below the FreeSurfer-segmented 5-mm subcortical band (Figs. 3C and 8), indicating that the pathological processes captured by these kurtosis-derived phenotypes are not confined to subcortical WM but extend into deeper WM.

In the kurtosis-positive WM phenotype, the *increase* in Kr with disease progression can be attributed to the *decrease* of the negatively contributing variation of the frequency distribution (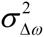 in Eq. [5]) to the kurtosis Kr. Indeed, the presence of frequency differences between axonal, myelin and extracellular water in WM is a signature of a rather regular cylindrical arrangement of bilayers forming lamellar myelin structure (Eqs. [11], [12]). The disease-induced variations in the cylindrical myelin lamellar arrangement, such as morphological changes in axons caused by axonal varicosities or beadings (*33, 34*), single-membrane herniations, myelin out-folds, and ectopic myelination, reported in AD mouse models (*35*) can reduce these frequency shifts, leading to increased metric Kr.

The situation is different in the kurtosis-negative WM phenotype where Kf becomes more negative with disease progression suggesting a decrease in R2t* relaxation variance and/or an increase in the frequency variation. Such changes in frequency distribution can hypothetically be caused due to the mesoscopic susceptibility effects (Eq. [9]) resulting from the presence of amyloid plaques (*36, 37*), iron (*38*), AD-induced changes in oxygen extraction fraction (*39*) and/or alterations in myelin structure (*40*) per previously suggested mechanism (*41*).

Previous work (*42*) has proposed a plausible pathway linking white matter structure to amyloid deposition involving cholesterol dysmetabolism as a general framework for white matter-amyloid interaction in AD: myelin injury releases cholesterol, which alters amyloid precursor protein processing and increases amyloid-beta production, while amyloid-beta in turn further damages myelin, forming a self-reinforcing cycle. The dual kurtosis phenotype described here may reflect two points along such a cycle - Kr indexing an earlier, more reversible structural axonal perturbation and Kf indexing a later, less reversible loss of myelin.

Subcortical white matter Kr changes were locally (defined by the FreeSurfer segmentation-matched WM-GM pairs) coupled to gray matter pathological changes and strengthened with disease stage. This pattern matches the anatomical signature of Wallerian degeneration, in which axons connected to a degenerating cortical region degenerate secondarily, and this effect intensifies as the upstream cortical pathology worsens. While this study quantifies cortical pathology via DM Fraction (a qGRE-related measure), the same Wallerian pattern - cortical pathology driving secondary, connected white matter degeneration that intensifies with disease severity - has been independently reported using tau deposition as the upstream driver (*43*), suggesting this mechanism may generalize across different AD pathological processes. This result is further compatible with postmortem evidence that white matter lesions in Alzheimer’s disease track cortical neurodegenerative pathology specifically, rather than small-vessel disease (*44*). Kf’s coupling, by contrast, was strong but essentially disease-stage-independent and comparable in matched and remote pairs, suggesting a stable, likely structural, relationship between local gray and white matter tissue properties that is present already in health rather than actively driven by disease.

This same distinction plausibly explains how Kr and Kf phenotypes relate differently to cognition. Cross-sectionally, both related comparably strongly to current cognitive performance. Longitudinally, Kr showed the strongest relationship to the subsequent, post-scan rate of decline; Kf also reached significance but contributed less. This pattern fits with what each parameter appears to reflect. Kf phenotype behaves like a largely static, structurally anchored property, so it should mainly track how much damage has already accumulated, thus informative about a person’s status, but less related to predicting where they are headed. Kr phenotype, by contrast, behaves like an active, disease-progressive process, consistent with its Wallerian-like coupling to gray matter pathology. Thus, parameter Kr should more directly capture the ongoing rate of neurodegeneration, which is consistent with its larger contribution to predicting future decline.

The diagnostic classification results (Fig. 5) reinforce this dissociation. Kr added substantial value to classifying clinical impairment beyond age, amyloid status, and APOE genotype, with a magnitude of contribution (27% of model performance in Shapley decomposition) second only to amyloid status itself (40%). Substituting each of the remaining three kurtosis-model parameters, or DM Fraction, a direct measure of gray matter pathology derived from the same qGRE acquisition, produced a graded pattern of diagnostic value (Kf > krR2t* ≈ kfR2t* ≈ DM Fraction) that mirrors the ordering seen throughout this study. That Kr outperforms DM Fraction specifically indicates this white matter signature is not merely a downstream readout of gray matter pathology status but carries independent classification-relevant information; together with its limited redundancy with amyloid and APOE genotype, this positions Kr as a genuine complement to, rather than a substitute for, established amyloid- and genetics-based predictors.

Including sex improved diagnostic accuracy by helping the model correctly interpret brain volume and Kr values, correcting the misclassification of two participants whose values were extreme outliers relative to their own sex but not relative to the sex-mixed cohort as a whole. This parallels findings for cognitive test norms in Alzheimer’s disease, where applying sex-specific rather than typical cut scores meaningfully improved diagnostic accuracy for amnestic mild cognitive impairment (*45*), indicating more broadly that a parameter’s relationship to sex-specific normative ranges, not only its raw value, can carry diagnostic information relevant to the design of future white matter biomarker models.

The myelin-structure-sensitive marker kfR2t* tracked Kf closely in both magnitude and behavior, whereas its positive counterpart, krR2t*, showed disease-stage progression comparable to Kr but did not share Kr’s anatomical coupling with gray matter pathology or its cognitive prognostic value. This dissociation is consistent with Kf reflecting more severe change accompanied by alterations in myelin structure, detectable by both the kurtosis-based Kf and kfR2t* measures. Kr, by contrast, appears to reflect a subtler structural change not necessarily accompanied by a variation in myelin content itself.

This study has several limitations. The cohort is drawn from a single research center, and while it spans the cognitively normal to mildly impaired spectrum with meaningful representation of both amyloid-positive and negative status, external replication in an independent cohort has not yet been performed. The biophysical interpretation of Kr as reflecting a rather subtle microstructural perturbation and Kf as reflecting more severe structural changes in WM, is grounded in the kurtosis model but has not been directly validated against histology in this cohort. Nine of 314 recruited participants who were clinically impaired but amyloid-negative were excluded from all analyses reported here, as this small, amyloid-discordant subgroup is unlikely to reflect AD-specific pathology and is too small to characterize reliably on its own. Most analyses here are cross-sectional; the longitudinal cognitive findings, while significant, are based on a reduced sample with variable follow-up duration and warrant confirmation in a larger, prospectively designed longitudinal cohort. Additionally, integrating tau PET, formal AD staging frameworks (e.g., A/T/N (*1*)), or vascular findings on MRI (e.g., white matter hyperintensities, microbleeds) with the RKI framework is an important direction for future work. Finally, Kr and Kf were each summarized as a single whole-cortex score; while this approach yielded robust, interpretable findings, it necessarily averages across regional heterogeneity that a more spatially resolved analysis might further clarify.

In conclusion, this work identifies a dual, sign-dissociable white matter kurtosis signature detectable within a single qGRE acquisition, requiring no additional scan time or contrast, that adds independent classification and prognostic value beyond standard clinical, genetic, and amyloid-based predictors, potentially filling the neurodegeneration WM component of the field’s dominant amyloid/tau GM biomarkers framework (*1, 46*).

## MATERIALS AND METHODS

### Study design

The study was approved by Washington University IRB. This was a single-center, cross-sectional and longitudinal observational cohort study designed to characterize a novel kurtosis-based extension of quantitative gradient recalled echo (qGRE) MRI relaxometry and to test its relationship to amyloid status, gray matter pathology, clinical impairment, and cognitive performance across the Alzheimer’s disease (AD) continuum. Participants were recruited through Washington University Knight Alzheimer Disease Research Center and underwent structural and qGRE MRI, amyloid biomarker ascertainment (positron emission tomography, PET, or cerebra-spinal fluid, CSF), APOE genotyping, clinical dementia staging, and neuropsychological assessment, with a subset followed longitudinally for cognitive outcomes. No participants were excluded based on imaging or biomarker results except as described below. Sample size was determined by availability of participants meeting inclusion criteria rather than by a prospective power calculation, consistent with a hypothesis-generating study of a novel imaging biomarker.

### Participants

The study cohort comprised 314 older adults spanning cognitively normal to mildly clinically impaired status (Clinical Dementia Rating, CDR, 0 to 1.0). Participants were classified into seven groups (Table 1) based on amyloid status (positive or negative, by PET or CSF), clinical impairment status (CDR 0, unimpaired, versus CDR >0, impaired), and APOE ε4 carrier status (carrier versus non-carrier). Amyloid-negative, clinically unimpaired participants (G1-G2) served as the healthy reference group; amyloid-positive, clinically unimpaired participants (G3-G4) represented a preclinical AD stage; and amyloid-positive, clinically impaired participants (G5-G6) represented a symptomatic AD stage. Within each of these three clinical strata, participants were further divided by APOE ε4 carrier status to allow the joint contributions of amyloid-related disease stage and genetic risk to be examined without confounding. A seventh group (G7) comprised participants who were clinically impaired but amyloid-negative. Because this amyloid-discordant presentation is unlikely to reflect AD-specific pathology and was too small to characterize reliably on its own, G7 was excluded from all primary analyses, leaving an analysis cohort of n=305 (G1-G6).

### MRI acquisition and images processing

Brain MRI was performed at Washington University in Saint Louis using 3T MRI (Siemens, Erlangen, Germany). The MRI protocol consisted of 3D multi gradient-recalled echo (mGRE) sequence with phase stabilization block and a standard clinical T1-weighted magnetization-prepared rapid gradient-echo imaging (MPRAGE).

MPRAGE parameters were: FOV = 256×256 mm, TI/TE/TR= 1100/3.37/2000 ms, flip angle= 10°, acquisition time 6 min, and resolution 1×1×1 mm^3^.

3D mGRE parameters were: FOV 256×192 mm, in-plane resolution 1×1 mm^2^ (read, phase), 80 2-mm slices, 10 gradient echoes with first echo time 4 ms, echo spacing 4 ms, repetition time 50 ms, flip angle 30°. A phase stabilization echo was collected at each TR to correct each line in k-space for the physiological-fluctuations-related phase inconsistancies (*47*). The total acquisition time with GRAPPA parallel imaging (*48*) was 6 min.

The acquired raw data of the mGRE sequences were reconstructed using in-house Matlab-based (MathWorks Inc., Natick, MA, USA) program. After phase correction for the physiological fluctuations (*47*) and GRAPPA reconstruction, k-space data from each receiver coil element were converted to the spatial domain subsequent to 2D in-plane Hanning filtering in order to reduce Gibb’s ringing and noise, and the resulting individual coil complex images were combined optimally using a previously developed algorithm (*49*).

Cortical and subcortical white matter and gray matter regions of interest were defined using FreeSurfer-based anatomical parcellation, co-registered to each participant’s qGRE data.

### Relaxation Kurtosis Imaging (RKI)

To account for the complicated multi-compartment distribution of water in the brain between a multitude of cells and extracellular environments, we describe the mGRE signal at a time TE after the radio frequency (RF) pulse excitation as a sum of exponentials with different tissue specific frequency shifts Δ*ω _j_*, tissue specific transverse relaxation rate parameters 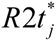, and the fractions *f_j_*:

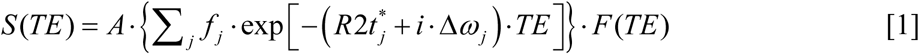

where *F(TE)* describes contribution of macroscopic field inhomogeneities to the GRE signal formation that is accounted for in the framework of the Voxel Spread Function (VSF) approach (*31*). Such a separation is possible due to the different spatial scales of interactions responsible for tissue specific (acting at the scale of molecules, cells and blood vessel network) and macroscopic field inhomogeneities that originate mostly from the susceptibility differences between brain tissue and surrounding air, acting at a scale of brain macrostructure (*50*).

In this paper we introduce an RKI approach that is based on a biophysical model approximating the signal Eq. [1] by a second order cumulant expansion:

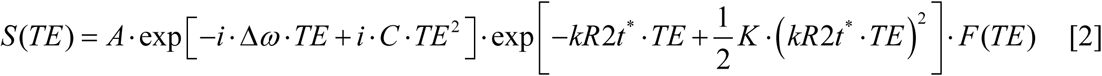

where kR2t* is the mean value of R2t* values across multiple water compartments *j* (the prefix “k” is added to distinguish the kurtosis-model-defined mean R2t* value from the R2t* value defined in the framework of the traditional qGRE biophysical model (*21*)):

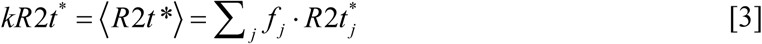

Δ*ω* is the mean value of the frequency shifts

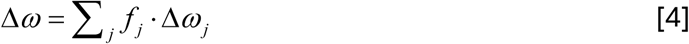

The cumulant-based relaxation kurtosis metric K (the second order cumulant expansion coefficient) has several contributions and is defined as follows:

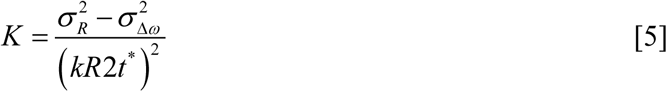

The Relaxation Variance 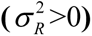 increases *K* due to the differences in compartmental decay rates:

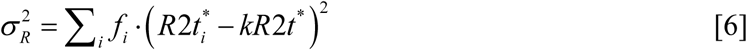

The Frequency Variance 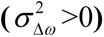 acts as a negative kurtosis *K* term due to the differences in the intercompartmental frequencies:

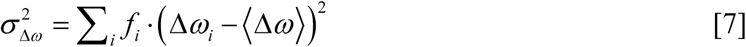

Parameter *C* in Eq. [2] is a relaxation-frequency covariance

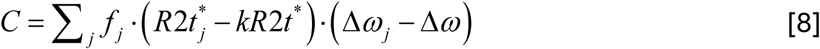

While the kR2t* relaxation is determined by both, the quantum spin interactions between protons in water and cell-building macromolecules, and the cellular level magnetic susceptibility effects, the frequency shifts are determined by the cellular level magnetic susceptibility effects (see for details (*51–55*)) and can also be affected by the additional mesoscopic (extracellular) magnetic susceptibility effects originating mostly from the sources such as blood vessels (i.e. BOLD), iron, etc. (often called magnetic susceptibility inclusions) that have magnetic susceptibility differences with the surrounding cellular environment (*30*). These effects will make additional contribution 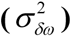 to the kurtosis (K) metric by modifying Eq. [5] as follows:

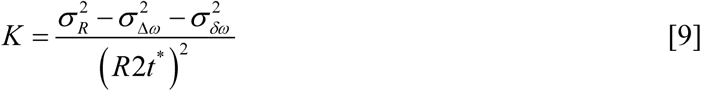

For a small volume fraction of magnetic susceptibility inclusions their contribution can be presented as follows (*30*):

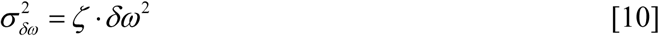

where *ζ* is the volume fraction of magnetic susceptibility inclusions and *δω* is the characteristic frequency difference between the magnetic susceptibility inclusions and the surrounding biological tissue. Note, that *δω* also depends on the shape and orientations of these inclusions with respect to the external magnetic field B0 (*30*).

For a commonly assumed three-compartment water model that includes axonal water, myelin water (water trapped inside the laminar structure of myelin) and extracellular water, the frequency shifts from the extracellular water were determined in (*54, 56*) as follows:

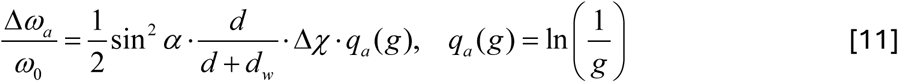

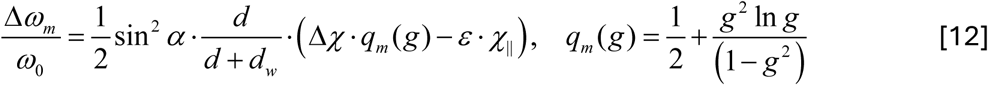

Here Δ*χ* = *χ*_||_ − *χ*_⊥_ is the difference between longitudinal and transverse components of magnetic susceptibility of lipoprotein chains in the myelin sheath, the g-factor is defined as *g* = *R_axon_* / *R_ext_*, *α* is the angle between the axonal and the external magnetic field **B**_0_ directions and *ε* defines the apparent fraction of time that a water molecule resides in the lipid layer. In fact, this parameter has a similar meaning as a parameter *n_w_* in the THB (transient hydrogen bond) model of MR signal relaxation (*57*).

The kurtosis model allows for a significant reduction of parameters characterizing the microstructure of biological tissues but should be applied with caution. The Eq. [2] provides an exact representation of Eq. [1] only for the Gaussian distribution of relaxation rates and frequencies. Otherwise, it is valid only for TE, smaller than the inverse of the characteristic differences (square root of variances) between inter-compartment relaxation times, and frequencies.

Note a formal mathematical analogy between RKI and diffusion kurtosis imaging (DKI) (*58*) – both account for quadratic terms in the signal (TE^2^ in RKI and b^2^ in DKI) – though having different biophysical origins.

### Gray Matter Pathology (Dark Matter)

In this paper we use the kurtosis model for analyzing qGRE signal in white matter. For analyzing the gray matter pathology we use the traditional qGRE biophysical model (*21*) characterizing each brain region by a single value of R2t*, a transverse relaxation rate specific to brain tissue itself, obtained after removing the contribution of the vascular BOLD effect (*30*) and magnetic field inhomogeneities (*31*). The GM regions with reduced R2t*, called dark matter (DM) (*27*), are considered pathological.

In this paper the DM status was defined relative to a healthy reference R2t* distribution built from a subset of participants with age <64 years, normal cognition, APOE ε4-negative, and cortical amyloid standardized uptake value ratio <1.42. For each FreeSurfer-defined cortical region, a pooled histogram of R2t* across these reference participants was constructed, and voxels with R2t* below (mean - 1.96×SD) of this reference distribution were classified as DM. DM Fraction for a given participant and region is the proportion of that region’s voxels meeting this criterion.

### R2t* vs. kR2t*

Note the difference between the notations: R2t* stands for the tissue-specific part of R2* after removal vascular BOLD and macroscopic magnetic field inhomogeneity effects as defined by the traditional biophysical model (*21*); kR2t*, on the other hand, stands for the tissue-specific part of the R2* relaxation rate parameter after removing effects of tissue microstructural heterogeneity (both, relaxation heterogeneity and frequency heterogeneity, including mesoscopic vascular BOLD effect) and macroscopic magnetic field inhomogeneities in the framework of the described above biophysical Kurtosis model, Eqs. [1]-[10]. We use R2t* metrics for analyzing the pathology of GM and kR2t* metrics for analyzing the pathology of WM.

In WM we further separate regions with positive, i.e. relaxation rate variation-dominated kurtosis, characterized by the Kr metric in conjunction with krR2t*; and regions with negative, i.e. frequency-variation dominated kurtosis, characterized by the Kf metric in conjunction with kfR2t*.

### PCA1-weighted whole-cortex parameter scores

For each of the four RKI model parameters (Kr, Kf, krR2t*, kfR2t*), a single whole-cortex score per participant was computed from the 32 established cortical white matter ROIs to provide a robust, low-dimensional summary measure for statistical analysis. Within the healthy reference group (G1, n=128), regional parameter values were standardized (z-scored) across the 32 ROIs, and principal component analysis (PCA) was applied to this standardized region-by-participant matrix. The first principal component (PC1) explained the majority of variance for each parameter (50.5% for Kr), with a sharp drop-off to under 10% for every subsequent component, indicating a single dominant factor underlying regional covariation. The resulting PC1 loadings were normalized to sum to 1 and applied as fixed weights to each participant’s raw (non-standardized) regional values, yielding one PCA1-weighted score per parameter per participant, used in all subsequent analyses.

### Amyloid status, APOE genotyping, and clinical assessment

Amyloid status was determined by amyloid PET (mean cortical standardized uptake value ratio [SUVR] ≥1.42, using Pittsburgh compound B) (*59, 60*) or, when PET was unavailable, cerebrospinal fluid (CSF) Aβ42/40 ratio (<0.0673) (*61*) as a fallback. APOE genotyping was performed using standard genotyping methods, and participants were classified as APOE ε4 carriers (one or two ε4 alleles) or non-carriers. Clinical impairment was assessed using the Clinical Dementia Rating® (CDR®) scale (*28, 29*), with CDR 0 defined as unimpaired and CDR >0 (0.5 to 1.0 in this cohort) defined as clinically impaired.

### Gray-white matter anatomical coupling

To assess disease-stage progression and gray-white matter coupling with adequate statistical power at the impaired stage (where the non-APOE ε4 carrier pathway includes only 9 participants), participants from both APOE ε4 pathways (carrier and non-carrier) were pooled at each of three disease stages (Unimpaired, Preclinical, and Impaired; Table 1). Each participant’s parameter value was first age-adjusted using their own pathway’s healthy reference group (G1 for the non-carrier pathway, G2 for the carrier pathway), then expressed as a deviation from that reference group’s mean, allowing participants from both pathways to be combined into pooled disease-stage groups without pathway-baseline differences confounding the pooled estimate. Coupling between each white matter parameter and gray matter pathology was computed as the correlation between each of 32 cortical gray matter regions’ DM Fraction and the corresponding white matter ROI’s parameter value (a matched pair), compared to the average correlation with all other, anatomically unrelated cortical regions (remote pairs), separately at each pooled disease stage.

### Classification of clinical impairment

Classification of clinical impairment (CDR-positive vs. CDR negative) was tested using repeated 10-fold cross-validated logistic regression (10 repeats) in the n=305 analysis cohort. Candidate predictors included age, amyloid status, APOE ε4 carrier status, sex, whole-brain volume (z-scored), and each PCA1-weighted kurtosis-model parameter or DM Fraction in turn. Model performance was quantified as the cross-validated area under the receiver operating characteristic curve (CV-AUC). For the best-performing model, the relative contribution of each predictor to overall cross-validated model performance was quantified using Shapley decomposition, a game-theoretic approach that attributes a model’s total performance additively across predictors by averaging each predictor’s marginal contribution across all possible predictor subsets (*62, 63*).

### Cognitive assessment

Current cognitive performance was assessed using the Global Composite score (*64*), derived from a standard neuropsychological test battery. Baseline cognitive assessments were matched to each participant’s MRI scan using the closest available assessment within a one-year window (mean interval 94.5 days, median 84.5 days, interquartile range 52-125 days). For participants with at least two cognitive assessments obtained after the MRI scan and spanning at least one year, a per-participant rate of subsequent cognitive change (points per year) was computed as the slope of a linear fit to these post-scan global composite score assessments (in the final analysis sample, n=128: mean follow-up span 3.3 years, median 2.4 years, interquartile range 1.4-4.6 years; median 3 assessments per participant, range 2-12). Relationships between each white matter or gray matter parameter and cognition were tested using joint linear regression models with age, amyloid status, APOE ε4 carrier status, and sex as simultaneous covariates, with either current global composite score or the subsequent rate of global composite change as the outcome; the relative contribution of each predictor was again quantified using Shapley decomposition of the model’s total R^2^.

### Statistical analysis

All statistical analyses were performed in Python (versions of NumPy, SciPy, pandas, and scikit-learn). Group comparisons of continuous parameters across disease stages used two-sided independent-samples t-tests; comparisons across the full disease-stage progression additionally used the endpoint (Unimpaired-vs-Impaired) contrast alongside adjacent-stage contrasts. Simple associations between parameters were assessed using Pearson correlation. Classification model performance was assessed using repeated k-fold cross-validation as described above, with predictor contributions quantified via Shapley decomposition. Statistical significance was set at P<0.05 (two-sided) throughout; exact P values are reported except where P<0.001. No correction for multiple comparisons was applied across the distinct analyses described here (disease-stage progression, gray-white matter coupling, diagnostic classification, and cognitive relationships), as each addresses a conceptually distinct question using a shared set of pre-specified kurtosis-model parameters; P values within each analysis should be interpreted in this context.

## FUNDING SOURCES

This study is supported by:

National Institutes of Health grant RF1/R01 AG077658 (DY)

National Institutes of Health grant RF1/R01 AG082030 (DY)

National Institutes of Health grant P30 AG066444 (JM)

National Institutes of Health grant P01 AG026276 (JM)

### Author contributions

Conceptualization: DY, SK

Methodology: DY, SK

Investigation: DY, SK, TB, CE, JM

Funding acquisition: DY, JM

Project administration: DY, SK, TB, JM

Supervision: DY, TB, JM

Writing – original draft: DY

Writing – review & editing: DY, SK, TB, CE, JM

### Data and materials availability

Data used in this paper are available from the [Knight Alzheimer Disease Research Center | Washington University in St. Louis](https://knightadrc.wustl.edu/) upon a reasonable request.

